# Sickle cell, thalassemia, and heat: risk estimates and equity policy considerations from an exploratory timeseries study in California

**DOI:** 10.1101/2024.10.15.24315547

**Authors:** Dharshani Pearson, Beth Apsel Winger, Keita Ebisu

## Abstract

**Background:** People born with hemoglobinopathies, such as sickle cell disease (SCD) and thalassemia, as well as people who are carriers for these disorders [sickle cell trait (SCT), thalassemia trait or thalassemia minor (TM)], may experience increased symptom-triggers and related illnesses from heat because their cells are susceptible to dehydration. Additionally, historical, and structural injustices could also exacerbate risk vulnerabilities among some communities.

**Methods:** In this work, we (1) present an overview of biological plausibility linking heat and hemoglobinopathy-related hospitalizations; (2) analyze the relationship between daily apparent temperature and such hospitalizations in California using Patient Discharge Data (PDD) and a two-stage timeseries study design with Poisson regression to estimate a state-wide hospitalization risk estimate; (3) discuss how structural barriers working in conjunction with demographic and genetic factors have conferred added risk to some Californians, thereby presenting deep-seeded equity ramifications.

**Results:** Our final dataset, restricted to the warm season, consisted of 96,908 daily counts of any hemoglobinopathy, listed as primary or secondary hospital diagnoses in PDD data. For an overall result, we observed an elevated risk of 3.0% (95% CI: 0.6, 5.5) per 10-degree Fahrenheit (°F) increase in apparent temperature at the last 30-day cumulative exposure window. For secondary hospital diagnoses of SCT-associated outcomes, when limited to very populous areas, we observed an elevated association of 7.0% at lag 23 (95% CI: 2.2, 12.1).

**Conclusions:** We observed excess symptom associations at various heat exposure levels, highlighting the need to examine longer exposure periods and potential care delays (stigma, medical racism, healthcare access). Differences and uncertainties could also stem from other comorbidities, age, genetics, housing and socioeconomic characteristics, or lifestyle variations. Education on the possible links between climate and hemoglobinopathy symptoms, changing demographics, and long overdue research funding may help close the gap in this interconnecting, critical environmental justice issue.

**Highlights:**

- People with hemoglobinopathies, or inherited red blood cell abnormalities, can experience health effects from high outdoor temperatures.
- Using over 20 years of hospitalization records, we examined a potential relationship among those who have an inherited hemoglobinopathy or those who are carriers.
- We found a connection between heat and hemoglobinopathy-related conditions over varying exposure periods lasting up to a month.
- Structural barriers to care and medical racism may exacerbate health outcomes during heat episodes for some of those who have inherited hemoglobinopathies, highlighting the need for novel strategies to combat disparities in healthcare access and quality of care.

## 1. Background and Purpose

Research suggests higher outdoor temperatures can increase hospitalizations and emergency room visits (ERVs) for heat illness, cardiovascular, and other related health events (Basu et al., 2012). Globally, researchers observe far-reaching vulnerabilities to heat among diverse populations (Lu et al., 2022). A recent review of existing studies concludes some comorbidities, including diabetes, pre-existing cardiovascular conditions, kidney disorders, and hypertension further increased risk of high temperature-related mortality and morbidity through sympathetic nervous system activation as well as inflammatory responses that involve multiple organ systems (Liu et al., 2015), supporting earlier research finding the same connections (Basu et al., 2012; Lu et al., 2022; Malig et al., 2019). Despite evidence for a strong relationship between heat and some broad disease categories, however, only a few researchers have delved into specific pre-existing or chronic health vulnerabilities among people seeking medical care following heat exposure (Hess et al., 2014).

Both hot and cold temperatures can influence hospitalizations depending on the specific disease. Therefore, focusing on specific chronic conditions can better characterize a temperature association. In this analysis, we narrowed our focus to hemoglobinopathies, which are inherited red blood cell conditions, because they can complicate or lead to comorbidities such as liver, thyroid, and kidney dysfunction, diabetes, hypertension, and infections (Kattamis et al., 2020; Kattamis et al., 2022; Weatherall, 2001), conditions that are also influenced by heat. Heat can exacerbate symptoms or related illnesses associated with inherited hemoglobinopathies.

In this work, we attempt five goals: (a) provide evidence and biological plausibility for an increased vulnerability, both directly and indirectly, to heat among those with hemoglobinopathies; (b) analyze the relationship between daily apparent temperature and hemoglobinopathy-related events in California using Patient Discharge Data (PDD) and a timeseries study design; (c) discuss how cumulative effects of genetic susceptibility, lack of disease knowledge, demographic shifts, and structural barriers predispose some populations in California to greater risk; (d) discuss how further research in this area can directly address the state’s commitment to a California for All, the California Strategic Growth Council’s Racial Equity Resolution and Equity Executive Order, EO N-16-22; (e) finally, discuss current and future public health implications in an increasingly warming climate and offer concrete mitigating strategies. In this study, “hospitalizations for hemoglobinopathies” refer to primary or secondary conditions that are directly related to being born with a hemoglobinopathy.

### 1.1 Genetics and biological plausibility

Sickle cell disease (SCD) and thalassemia are autosomal recessive inherited hemoglobinopathies, meaning that typically, only disease-conveying genes from both parents result in full disease manifestation (Figures 1 and 2). SCD results in sickle-shaped red blood cells instead of the standard flexible, donut-shaped healthy red blood cells. These sickle cells are hard and lack the flexibility of normal cells, a characteristic that leads cells to clump together, clot blood vessels, and cause a painful condition called vaso-occulusion or sickle cell pain episodes, which can last days and even weeks (UK, 2022). Triggers such as dehydration, certain foods, infections, and extreme external temperature (either hot or cold) can set a pain episodes in motion as blood consistency (including viscosity) changes (Ochocinski et al., 2020; Smith et al., 2003; Tripette et al., 2013). In the United States (U.S.), SCD disproportionately impacts Black Americans: in 2010, the incidence of SCD was 73.1, 6.9 and 3.0 cases per 1,000 Black, Hispanic, and White births, respectively (Ojodu et al., 2014). Latinx SCD births currently represent 10% of California newborns diagnosed with SCD, exhibiting a clear increase across groups (Valle et al., 2022). Although newborn screening for SCD and sickle cell trait (SCT, one copy of a mutated hemoglobin gene) is mandatory in the U.S., many with SCT are unaware or unsure of being a carrier, especially if they are older or were born in a different country (Bean et al., 2014). Despite the reporting requirement, states differ in test administration and how patients receive results and explanations (Kavanagh et al., 2008). Although California currently conducts robust surveillance, older Californians or those born in other states may still lack proper diagnosis.

**Figure 1.**
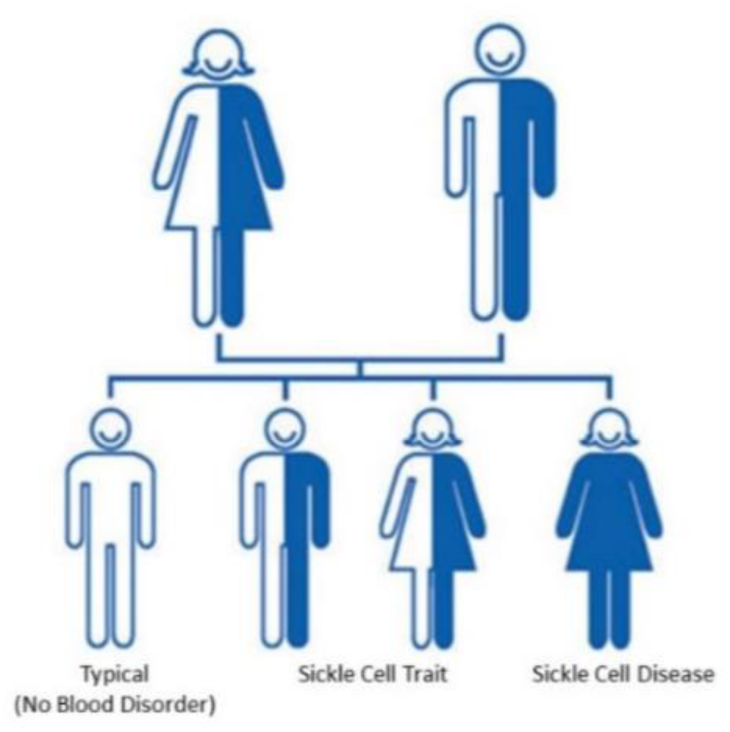
Sickle Cell Trait Inheritance Materials developed by CDC. Reference to specific commercial products, manufacturers, companies, or trademarks does not constitute its endorsement or recommendation by the U.S. Government, Department of Health and Human Services, or Centers for Disease Control and Prevention. This material is available at the CDC website at no cost. https://www.cdc.gov/ncbddd/sicklecell/traits.html

**Figure 2a.**
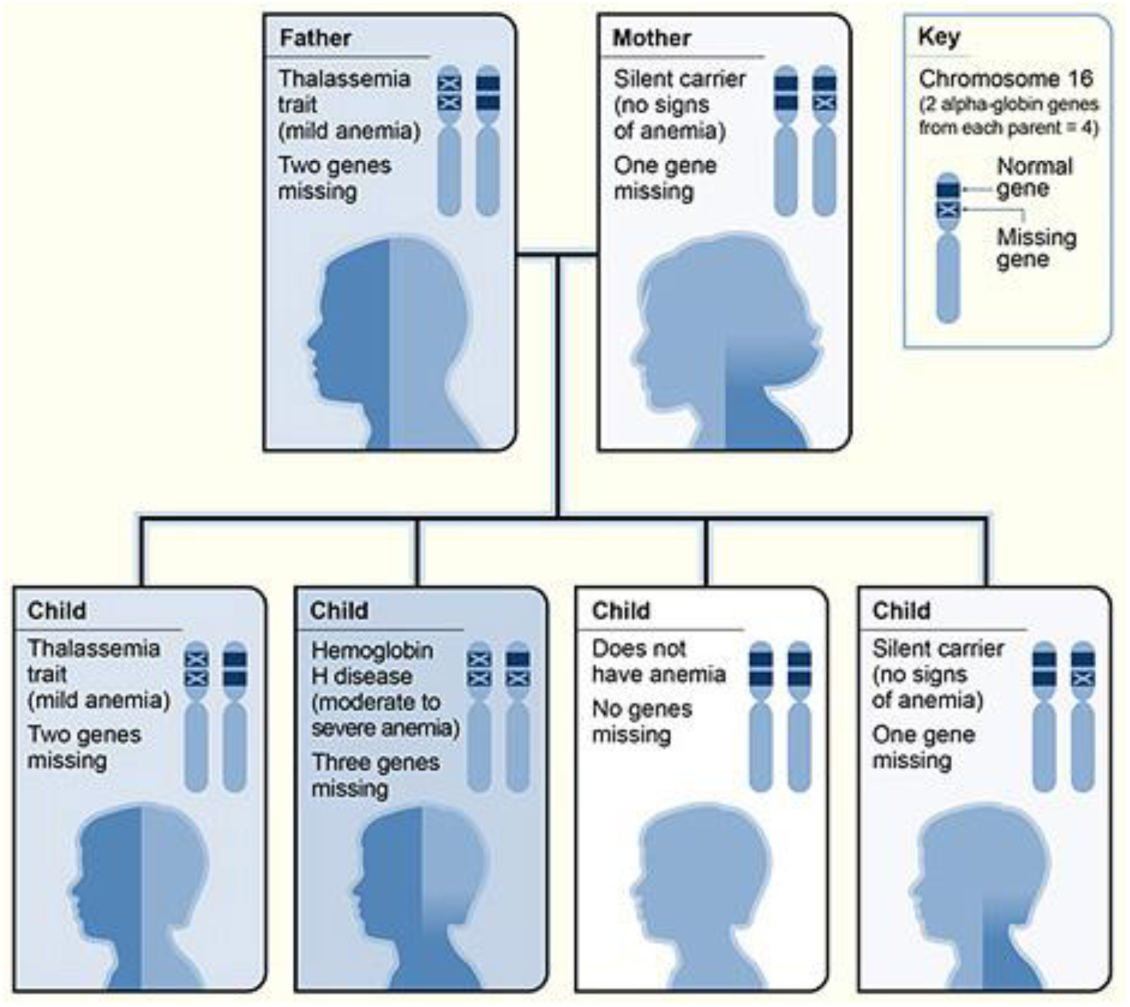
Inheritance Pattern for Alpha Thalassemia Source: National Heart, Lung, and Blood Institute; National Institutes of Health; U.S. Department of Health and Human Services. Reference to specific commercial products, manufacturers, companies, or trademarks does not constitute its endorsement or recommendation by the U.S. Government, National Heart, Lung, and Blood Institute, National Institutes of Health, or U.S. Department of Health and Human Services. This material is available at no cost here: https://www.nhlbi.nih.gov/health/thalassemia/causes

**Figure 2b.**
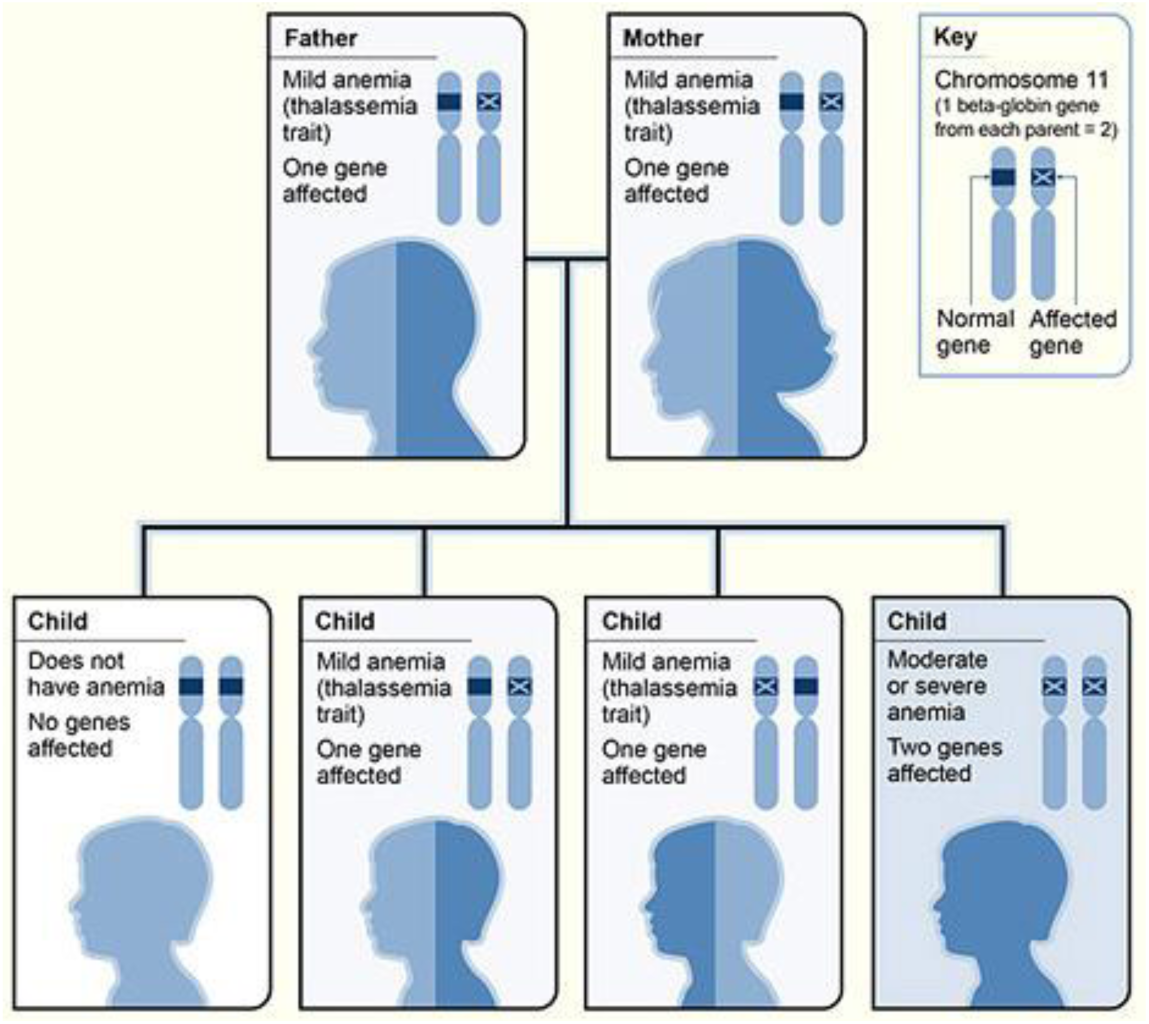
Inheritance Pattern for Beta Thalassemia Source: National Heart, Lung, and Blood Institute; National Institutes of Health; U.S. Department of Health and Human Services. Reference to specific commercial products, manufacturers, companies, or trademarks does not constitute its endorsement or recommendation by the U.S. Government, National Heart, Lung, and Blood Institute, National Institutes of Health, or U.S. Department of Health and Human Services. This material is available at no cost here: https://www.nhlbi.nih.gov/health/thalassemia/causes

Unlike SCD, thalassemia results from mutations in multiple genes coding for alpha and beta subunits of the hemoglobin protein, which involves oxygen binding in red blood cells. A mutation resulting in a non-functional alpha subunit is called alpha thalassemia (αT) while a mutation leading to a non-functional beta subunit is called beta (βT) thalassemia. Thalassemia has a wide range of health impacts, from benign to severe conditions incompatible with life. Patients who have severe forms of thalassemia need chronic or periodic blood transfusions throughout their lives (Lal and Vichinsky, 2023; Lal et al., 2021; Vichinsky et al., 2018). The Centers for Disease Control and Prevention (CDC) and National Institutes of Health’s National Heart, Lung, and Blood Institute (NHLBI) provide more information on both hemoglobinopathies and inheritance patterns (CDC, 2023a; CDC, 2023b; NHLBI, 2023).

Those who inherit one copy of a mutated hemoglobin gene are carriers: SCT carriers, or in the case of thalassemia, thalassemia trait carriers, or thalassemia minor (TM). Generally, people who are carriers for either disorder do not have symptoms; however, symptoms can occur, especially under strenuous exercise or dehydration associated with SCT (Reeves et al., 2019). Though still debated, both anecdotal evidence from practitioners and research increasingly indicate SCT is not a benign condition, with one study showing a statistically significant increase for direct SCT hospital admissions for children from 2006 to 2015 (Peterson et al., 2020; Udechukwu, 2021). Additionally, symptom severity among carriers can vary, sometimes depending on co-inheritance of another hemoglobinopathy, comorbidity, or other factors (Lee et al., 2022; Raffield et al., 2018). SCT carriers can have complications in high altitudes, while scuba diving, or while receiving anesthesia, and may also face kidney and liver complications, retinopathy, venous thromboembolic events, pregnancy complications, excessive urinary tract infections, and, rarely, kidney cancer (Hulsizer et al., 2022; Reeves et al., 2019; Tsaras et al., 2009). Some SCT carriers experience a delayed diagnosis of diabetes due to differences in how blood cells among those with SCT respond to hemoglobin A1c (HbA1c) pre-diabetes testing. (Lacy et al., 2017). In this instance, those carriers may have uncontrolled diabetes for longer, sustain more organ damage, and experience more complications (Lacy et al., 2017). There is also similar evidence, though more varied, with HbA1c and TM (Mitchai et al., 2021). Those with TM may rarely have symptoms related to anemia, but some research suggests that comorbidities along with TM can lead to further health complications (Lee et al., 2022); however, in-depth research into SCT or TM interactions with various exposures is lacking.

Previous studies show people with inherited blood disorders, specifically SCD and, to a lesser extent, thalassemia, manifest symptoms due to extreme external temperature, extreme temperature fluctuation, and/or season (Borgna-Pignatti et al., 2006; Brandow et al., 2019; Brandow et al., 2013; Smith et al., 2009; Smith et al., 2003; Wachnian et al., 2020). These symptoms can result from blood cells responding to heat and cold by sickling (Brandow et al., 2013; Smith et al., 2003). Biologically, *in vitro* evidence has demonstrated temperature changes, along with other factors, can influence both how much oxygen binds to hemoglobin as well as how often sickling takes place (Bohr, 1904; Eaton, 1994; Smith et al., 2003). A lower percentage of oxygen saturation can affect red blood cell rigidity, one component that can lead to vaso-occlusion (Mackie and Hochmuth, 1990; Smith et al., 2003). Results from one *in vitro* analysis indicated that at 37°C (typical body temperature), cell rigidity among sickled cells was 18 times that of normal cells (Mackie and Hochmuth, 1990). Less discussed in the literature is how thermal sensitivity could occur from secondary, related chronic conditions that may result from an inherited blood disorder condition, including kidney damage, deep vein thrombosis, diabetes, or thyroid issues. This sensitivity in people with SCD is unsurprising; however, climate change could cause more frequent extreme heat events, meaning SCT carriers, who typically experience few symptoms under normal circumstances, may experience heat stress more frequently and, over years, develop greater susceptibility to heat effects.

### 1.2 Existing Literature and Research Gaps

Research linking temperature changes and blood disorders, particularly SCD, is not new. Earlier studies have identified colder weather or winter season with increased risk of hospital or EDVs (Almuqamam et al., 2021; Ibrahim, 1980; Redwood et al., 1976; Rogovik et al., 2011; Xu and Frenette, 2019). Others have identified risk at both extreme temperature spectrums (Mekontso Dessap et al., 2014; Smith et al., 2003).

Most literature on SCT and environmental factors has focused on exertional heat stress and more severe outcomes, including death, especially among youth athletes as well as military personnel engaging in heavy training (Nelson et al., 2018; Singer et al., 2018; Tripette et al., 2013). In SCT, some conditions can promote sickling during strenuous exercise, leading to severe oxygen deprivation (hypoxemia), metabolic acidosis, muscular hyperthermia, and red-cell dehydration ((NATA), 2007).

In contrast, little information exists on non-fatal, yet serious heat and exertion-related health outcomes related to SCT that interfere with daily activities of living and quality of life. Media attention and research have focused on the worst outcomes of exertional-related SCT complications: those pertaining to mortality, which are rare. However, general studies on heat effects show associations with morbidity as opposed to just mortality. The same associations may stand true for SCT-linked hospitalizations as opposed to mortality. Thus, although exertional sickling and death may be rare, other conditions, like dehydration or syncopal incidents, could result from heat-SCT relationships.

Complicating matters further, medical staff could record hospitalizations under other diagnoses without mentioning SCT, thereby underestimating a potential link with a pre-existing condition. Because over 7% of Black Americans and nearly 1% of Latinx Americans carry SCT, these unrecorded or missing information visits could underestimate the burden of disease attributed to heat-SCT health outcomes for specific communities (Ojodu et al., 2014).

Symptoms of SCT can vary from person-to-person, so identifying higher risk groups could more efficiently allocate funds for the most vulnerable carriers. For instance, some evidence suggests co-inheritance of an α-thalassemia mutation (-α3.7 deletion) significantly lowered the risk of anemia and chronic kidney disease among Black Americans who also had SCT (Raffield et al., 2018), so lacking this co-inheritance could mean more risk for some SCT carriers.

Given the plausible biological connection to heat exposure and disproportionate health impacts to already impacted vulnerable populations, a state-wide analysis on the topic could better connect the multifactorial links between heat, hemoglobinopathies, and environmental justice issues (Figure 3).

**Figure 3.**
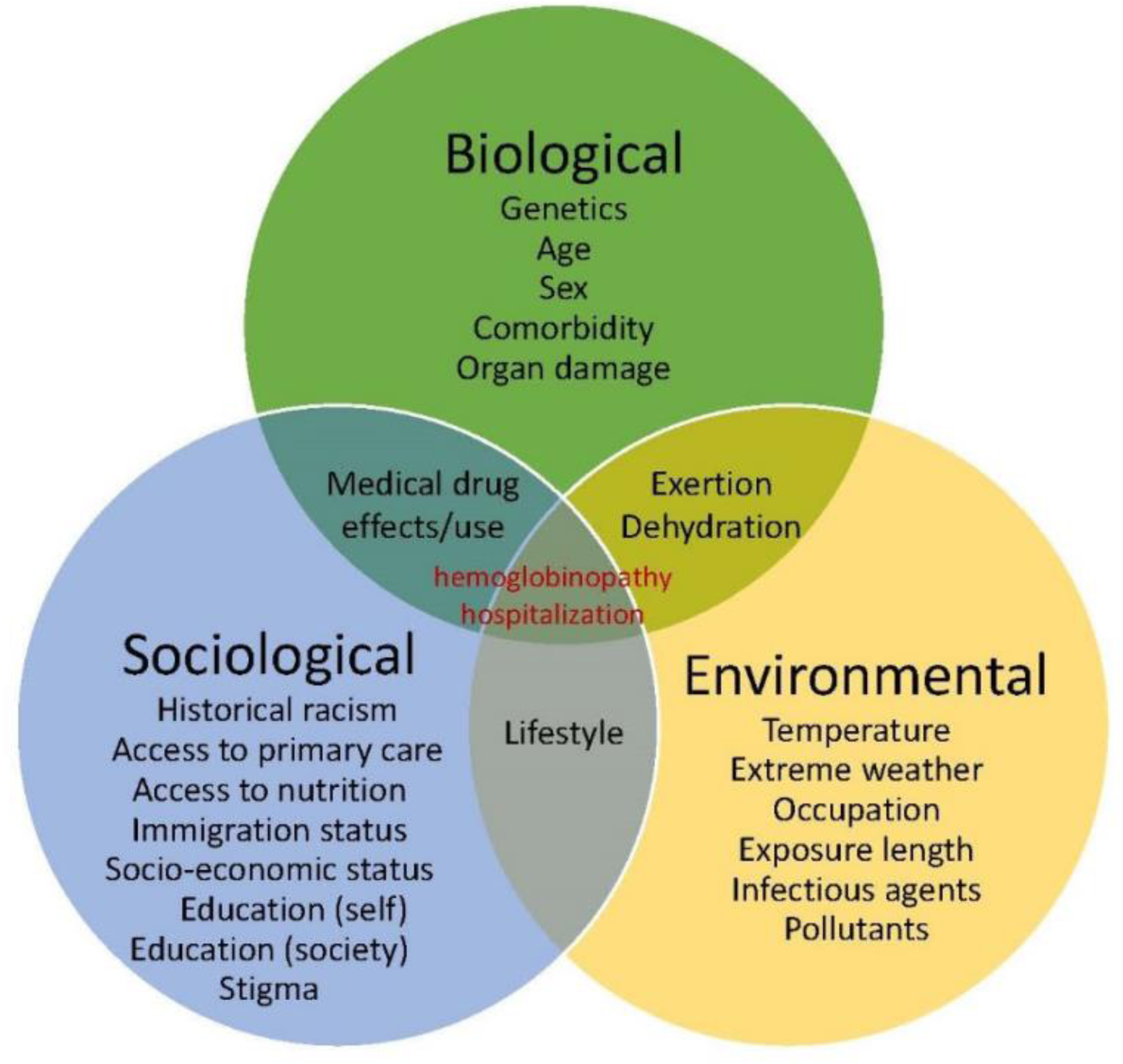
Interactions leading to hemoglobinopathy-related hospitalizations

## 2. Methods

### 2.1 Study Population

The California Department of Health Care Access and Information (HCAI) provided PDD for daily hospitalization counts of various categories of hemoglobinopathies, designated in the International Classification of Diseases, Ninth Revision (ICD-9-CM) codes 282.4 to 282.69, inclusive and Tenth Revision (ICD-10-CM) codes D56-D57, inclusive, from January 1999 through December 2019. PDD include all visits that lasted longer than 24 hours at a hospital. We could not exclude repeat visits, and multiple visits may have occurred for the same person. Although we used visits labeled with ICD codes spanning two revisions (switching from ICD-9-CM to ICD-10-CM in October 2014), the pattern of utilization did not change markedly between the coding schemes. We looked at both primary and all secondary coded diagnoses outcomes (up to 24) for a hemoglobinopathy. A primary diagnosis means a person with a hemoglobinopathy was admitted for an hemoglobinopathy-specific symptom, such as for pain crisis in Primary SCD. For all secondary hemoglobinopathy diagnoses categories, a person was admitted for a primary cause other than a hemoglobinopathy, such as dehydration or a cardiovascular event, but a hemoglobinopathy was considered a contributing factor in the admission.

We obtained daily PDD counts for each of California’s 16 climate zones (CZs), by summing daily PDD counts in all ZIP-code tabulation area (ZCTA) within the CZs, which are California Energy Commission-defined regions based on weather and energy use factors (Figure 4) (Commission, 1995). ZCTA is based on residential ZIP code and is a variable available in PDD.

**Figure 4.**
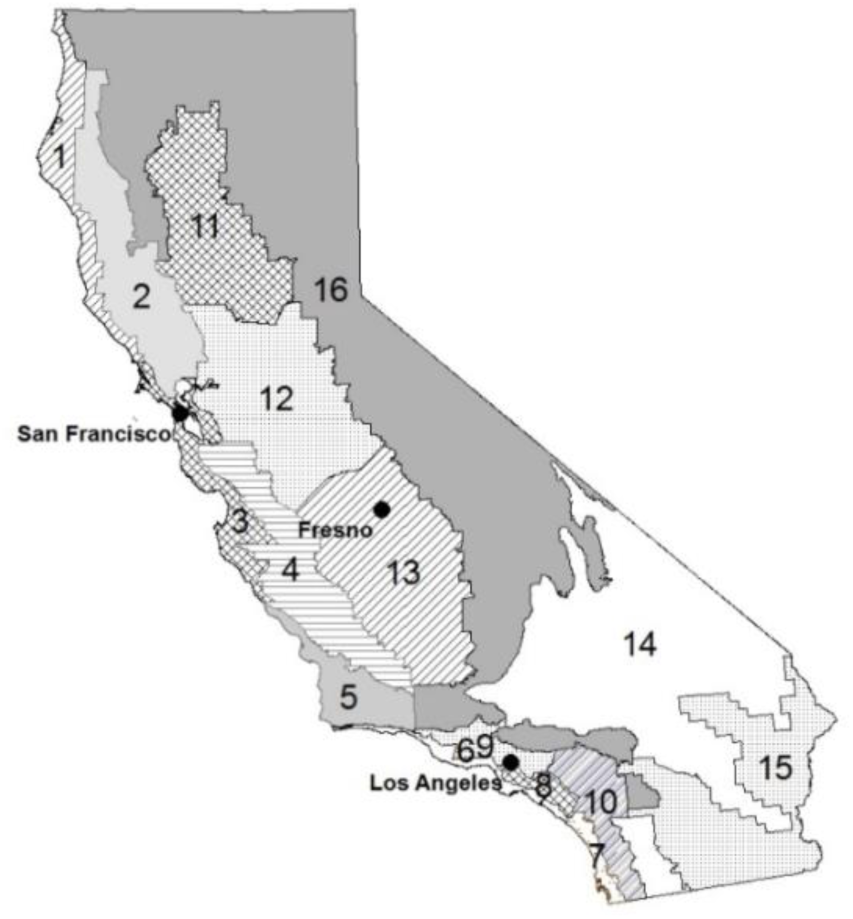
State of California with 16 Climate Zones

### 2.2 Exposure Data

We obtained daily temperature and relative humidity data from US EPA Data Mart (Agency, 2014), the California Irrigation Management Information System (Office of Water Use Efficiency, 2014), and the National Oceanic Atmospheric Administration (NOAA, 2012). The exposure dataset ranged from January 1999 to end of December 2019. We only utilized weather data where monitors had at least 95% completeness based on daily observations. We focused on daily apparent temperature, which considers both temperature and relative humidity. Because we were interested in heat associations, we only examined PDD visits from California’s warm season: May through October.

We calculated daily mean apparent temperature for each of California’s 16 CZs. First, we estimated the ZCTA-specific daily apparent temperature values using inverse distance weighting from monitors to the population-weighted centroid of the ZCTA within the same CZ. Earlier epidemiologic studies utilized this climate classification method to explore the relationships between temperature and health (Basu et al., 2018; Pearson et al., 2020). We then estimated mean apparent temperature values of each CZ by taking the population weighted average of values from all the ZCTAs within the same CZ. The 2010 U.S. Census Data provided population information. We used R Version 4.1.2 (R Core Team, 2021) for distance calculations, with additional data manipulation and cleaning completed using SAS Version 9.4 (Inc., 2014).

### 2.3 Data Analysis

We applied a two-staged approach for data analysis. In the first, we calculated effect estimates for each CZ using a time-series method with Poisson regression in R. In addition to daily mean apparent temperature, we added a natural spline smoothing function with a sequential day within each year to adjust for seasonal trend as well as indicator terms for weekend, holiday, and year. Then, in the second step, we combined all CZ results using random effects models for meta-analyses to obtain an overall estimate (Basu and Malig, 2011).

Because of the range of conditions examined, we assumed people may experience symptoms over a varying period of days. Thus, we decided to examine several exposure windows, also called lag periods, of apparent temperature to set the groundwork for future investigations. For this study, we considered lags of 30 single days or under and various combinations of multiple day averages of apparent temperature across the 30-day exposure window. Lag 0 represents same-day exposure as the date of hospitalization and Lag 1 represents the previous day exposure, for example.

We obtained Institutional Review Board (IRB) approval from California’s Committee for the Protection of Human Subjects (CPHS) prior to starting this study. We report all results as an excess percent increase in PDD visits for each 10-degree Fahrenheit (°F) increase in apparent temperature.

## 3. Results

Table 1 summarizes mean daily apparent temperature during the warm months for all of California and the 16 CZs. During the study period, the average warm month apparent temperature was 67.2°F for all CZs. Our final dataset consisted of 96,908 daily counts of any hemoglobinopathy, designated as a primary or secondary diagnosis. Table 2 illustrates more descriptive statistics by primary or secondary diagnosis and breakdown by major category (SCD, SCT, thalassemia major, TM), all primary hemoglobinopathy, or all secondary hemoglobinopathy diagnoses according to ICD code.

**Table 1.**
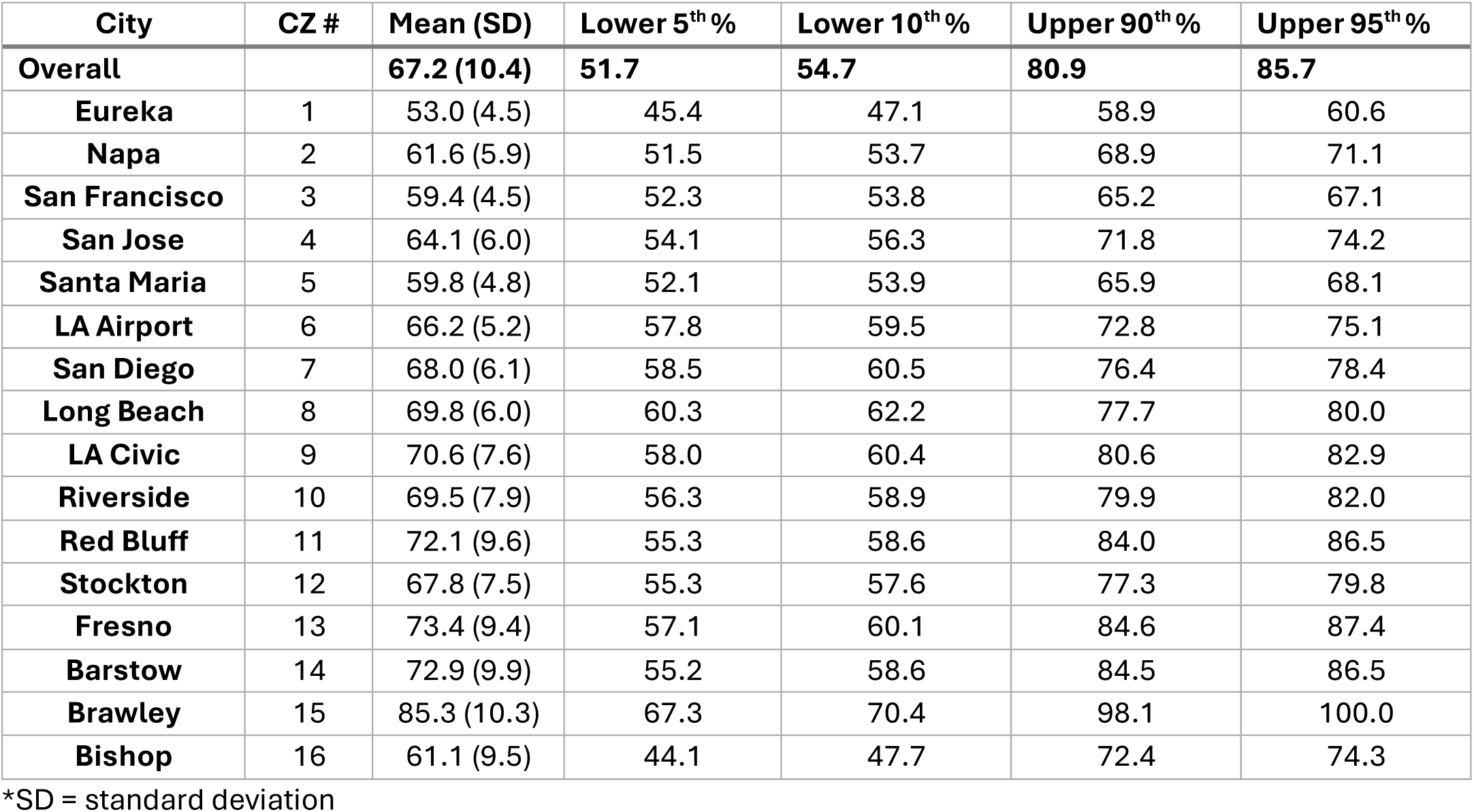
Mean apparent temperature exposure (°F), Standard Deviation (SD), and Select Quantiles by climate zone (CZ) with representative city for Warm Season (May-October) in California, 1999-2019.

**Table 2.** Descriptive Characteristics of the Study Population (%) in California, 2005-2013*.

| Outcome Category | ICD-9 | ICD-10 | Diagnosis Type | N (%) |
| --- | --- | --- | --- | --- |
| **All Hemoglobinopathies** | 282.4-282.69 | D56-D57 | Both | 96908 |
| **All Primary** | 282.4-282.69 | D56-D57 | Primary | 49171 (50.7) |
| **All Secondary** | 282.4-282.69 | D56-D57 | Secondary | 47737 (49.3) |
| **Primary SCD** | 282.6-282.69 | D57-D57.29;<br>D57.80- D57.819 | Primary | 45769 (47.2) |
| **Secondary SCD** | 282.6-282.69 | D57-D57.29;<br>D57.80- D57.819 | Secondary | 15722 (16.2) |
| **Secondary SCT** | 282.5 | D57.3 | Secondary | 9305 (9.6) |
| **Thalassemia** | 282.4, 282.43-282.45;<br>282.47-282.49 | D56-D56.29<br>D56.4 to D56.9 | Both, mostly<br>Secondary | 14736 (15.2) |
| **Thalassemia Minor** | 282.46 | D56.3 | Both, mostly<br>Secondary | 5057 (5.2) |

\* Categories are not mutually exclusive, except for All Primary and All Secondary.

**Table 3.** Selected Lag Results per 10°F increase in mean apparent temperature during warm season in California, 1999-2019.

| Outcome | Lag | % change (95% CI ) |
| --- | --- | --- |
| All Hemoglobinopathies | 0_30 | 3.5 (0.9, 6.2) |
| SCD (Primary) | 28 | 1.9 (0.1, 3.8) |
| SCD (Primary) | 0_30 | 3.6 (-2.3, 9.8) |
| SCD (Secondary) | 1 | 4.2 (-0.6, 9.3) |
| SCT (Secondary) | 23 | 5.5 (1.4, 9.8) |
| SCT (CZs 3, 8-10, 12) | 23 | 7.0 (2.2, 12.1) |
| All Thalassemia | 0_30 | 7.1 (-0.2, 15.0) |
| Cardiovascular Primary* | 27 | 9.1 (0.0, 18.9) |

CI, confidence interval
\*Did not include thalassemia major. Very small numbers and should be interpreted with caution

Looking at 30 days of exposure, for an overall result encompassing any hemoglobinopathy diagnosis code (primary and secondary), we observed an elevated risk of 3.0% (95% confidence interval (CI); 0.6, 5.5%) at Cumulative Lag 0_30, or 30-day average. We saw a lagged and cumulative effect for any hemoglobinopathy coded as primary diagnoses (Figure 5). SCD accounted for nearly all counts in this analysis. We observed increased trend from Lag 19 to Lag 28. Particularly, at Lags 19 and 20 and then again at Lags 26-28, we observed increased risk of about 2 percent in hospitalizations for a primary diagnosis. Additionally, when examining cumulative lags, we saw a greater increased risk trend as cumulative exposure lags increased, reaching a peak at Cumulative Lag 0_27 (3.6%; -1.1,8.6%) and Cumulative Lag 0_30 (4.0%; -1.0, 9.3%).

**Figure 5.**
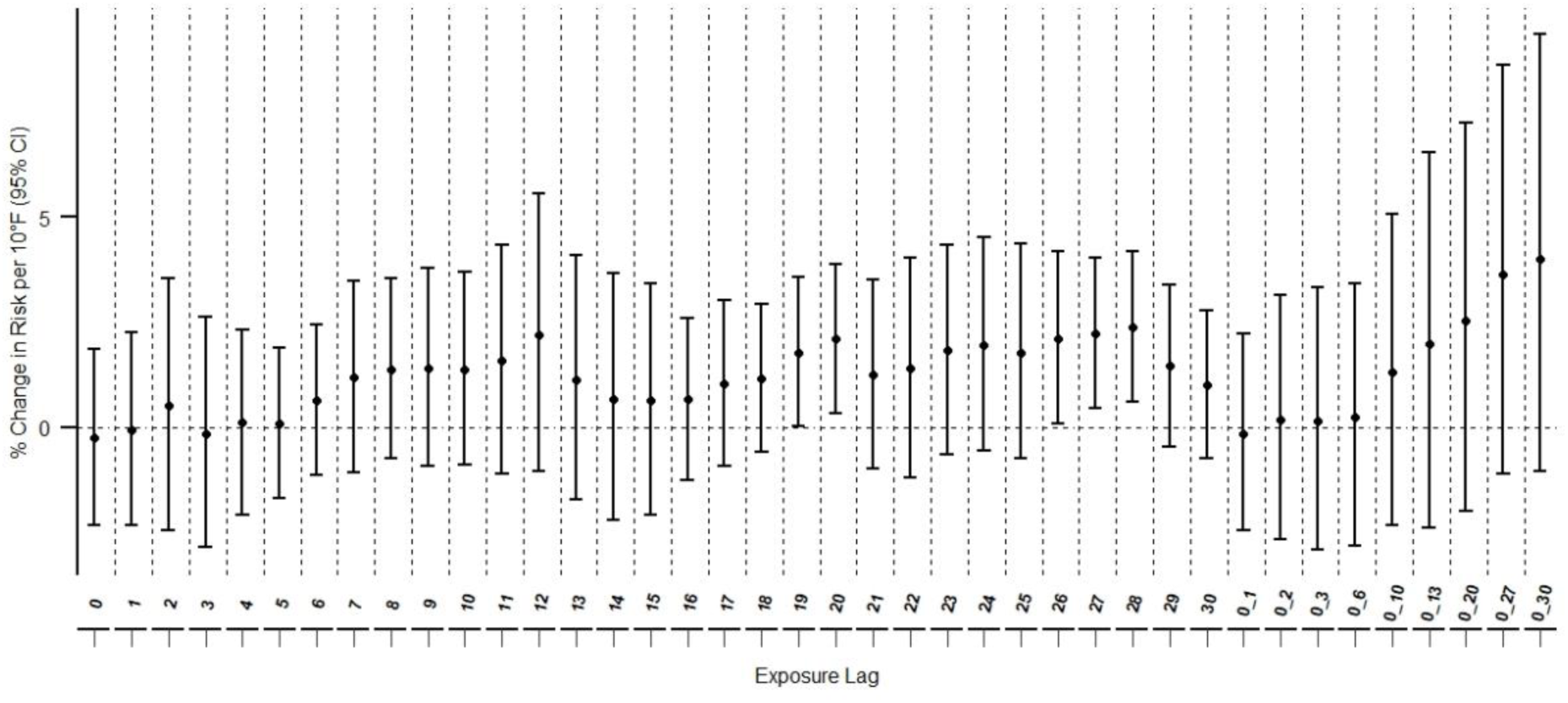
Risk of hospitalization for any primary hemoglobinopathy outcome (lags 0 to 30, selected cumulative lags 0_1 to 0_30)

In examining all secondary diagnoses of a hemoglobinopathy, meaning admittance for a non-hemoglobinopathy primary diagnosis, we observed an elevated immediate risk at Lag 1 of 2.4% (0.6, 4.3%), and then an elevated risk at Cumulative Lags 0_1 of 2.3% (0.4, 4.3%), 0_2 of 2.3% (0.3, 4.3%), and 0_3 of 2.2% (0.1, 4.3%). We also detected an increased risk at Lag 23 of 2.0% (0.0, 3.9%) (Figure 6).

**Figure 6.**
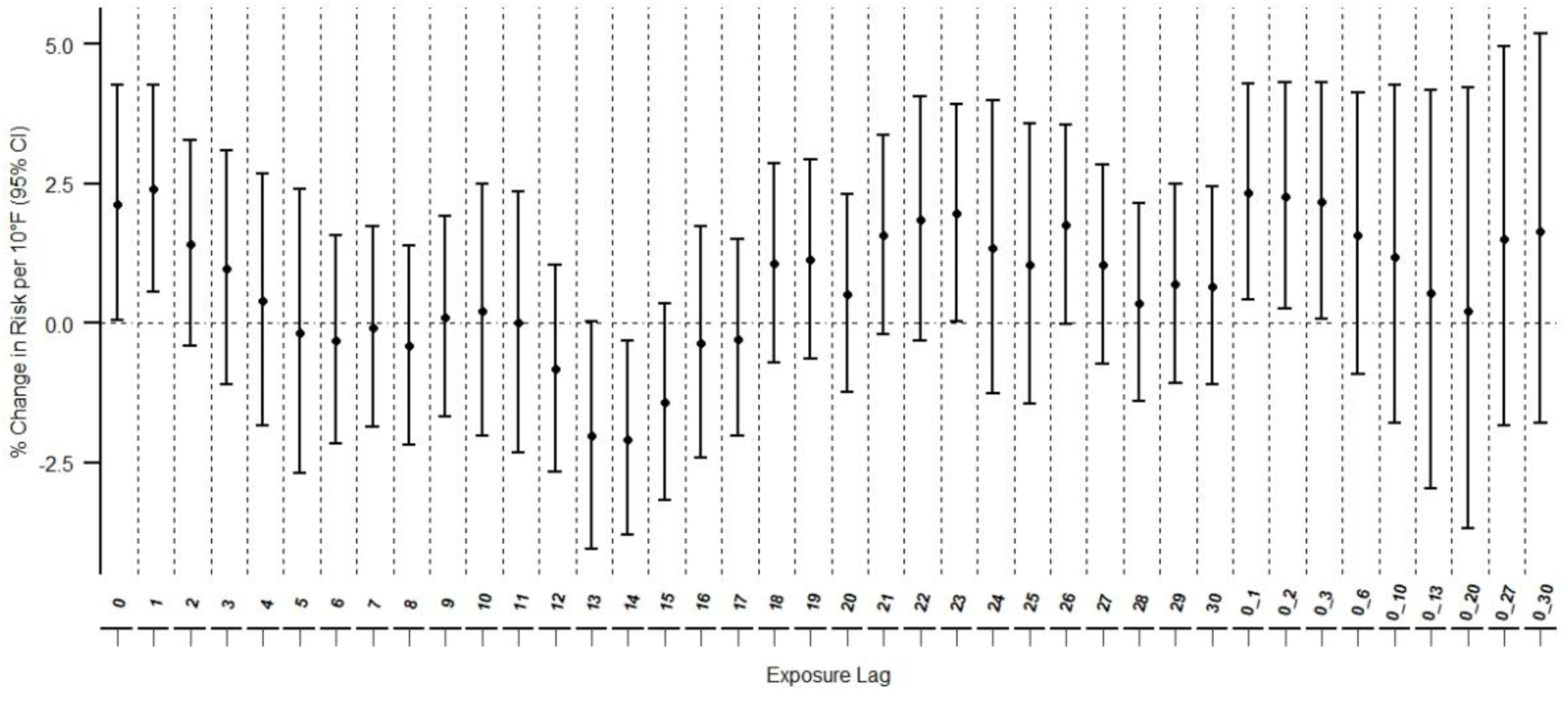
Risk of hospitalization for any secondary hemoglobinopathy outcome

Despite small numbers, we tried to look at SCD and SCT separately. For SCD, limited to primary diagnoses, we did not see any meaningfully different results from the all-hemoglobinopathy primary analysis, though smaller in magnitude for Lag 28 (1.9%; 0.1, 3.8%) and Cumulative Lag 0_30 (3.6%; -2.3, 9.8%). For SCD limited to secondary diagnoses, however, we detected an acute excess risk trend particularly for Lag 1 (4.2%; -0.6, 9.3%) and Cumulative Lag 0_1 (4.4%; -1.1, 10.2%). For SCT secondary diagnoses, on the other hand, we observed delayed elevated risk a little over three weeks later at Lags 22 (4.0%; -0.1, 8.2%), 23 (5.5%; 1.4, 9.8%), and 24 (4.5%; 0.4, 8.7%). When we examined the same outcome, limited to CZs 3, 8, 9, 10, and 12, the zones with the highest populations with SCT, we observed an even greater association at Lags 22 (5.0%; 0.3, 10.0%), 23 (7.0%; 2.2, 12.1%), and 24 (6.4%; 1.1, 12.1%).

We also attempted an analysis examining all thalassemia outcomes. In this analysis, although we observed increases throughout the 30 days, we again saw a pattern of cumulative effects as well as a small elevation at single Lags 22 to 30, particularly strong between Lags 22 to 24. At Cumulative Lag 0_30, we observed a 7.1% (-0.2, 15.0%) trend increase in risk among those with any thalassemia diagnosis. Additionally, we detected a smaller, though with narrower CIs, elevated risk trend at Lag 1 of 3.1% (0.5, 5.8%) and at Cumulative Lag 0_3 of 3.5% (0.1, 7.0%), indicating a more immediate effect.

Our final analysis explored primary cause categories where a hemoglobinopathy was recorded as a secondary diagnosis. We excluded thalassemia major from the secondary group because this group historically exhibited more uncertainty in diagnosis coding accuracy. Because of low daily counts, we could only examine broad categories of cardiovascular outcomes. For all primary cardiovascular events with a secondary hemoglobinopathy code, results showed a strong increasing risk trend during the third week after exposure, with Lag 27 showing the strongest association at 9.1% (0.0, 18.9%) when limited to CZs with the largest populations (CZ: 3, 8, 9, 10, 12).

## 4. Discussion

Along with discussing results, we explore biological, structural, and social components that in conjunction may explain some of our findings.

### 4.1 Implications from study results

Earlier studies looking at similar relationships had focused on very short-term associations, targeting under one week of exposure, showing mixed results (Smith et al., 2003). In our study, we saw a small association between higher apparent temperature and both primary and secondary hemoglobinopathy-related hospitalizations. We saw a delayed and cumulative effect related to heat and hospitalizations for a primary diagnosis, which were mainly SCD, showing the possible build-up to a pain episode days after exposure. Alternatively, people may also delay seeking care at hospitals due to stigma, fear of mistreatment, and disbelief of their pain severity in seeking pain relief medication, often prescription opioids. Some literature has described this fear, which prolongs suffering among those experiencing a pain episode (Farooq et al., 2020; Nogrady, 2021). Access to care, discussed further below, could also play a role in the delay. In contrast, when we examined all secondary outcomes, we observed both a strong immediate effect and lagged effect, depending on the outcome. For people with SCD secondary diagnoses, we detected a strong immediate effect, perhaps indicating a vulnerability among those with a blood disorder to primary conditions with acute responses to heat, including heat illness or heat stroke.

When looking at just secondary hospital diagnoses of SCT, we saw a more elevated association about three-and-a-half weeks after exposure. This association could stem from a primary hospital diagnosis taking longer to develop, such as with venous thromboembolism (mainly pulmonary emboli), which has been observed in SCT carriers and can manifest quite mildly at first and then worsen over time (Kato, 2019; National Heart, 2022). Though with small numbers with a wider CI, we did see an elevated 9% primary cardiovascular risk with a secondary blood disorder at about lag 27. For comparison, a California case-crossover study on hospitalizations from 1999-2005, which by design only examined exposures less than one week, showed a below 2% risk for cardiovascular outcomes at a much earlier lag, though we are cautious with this comparison because of study period and methodological differences (Ostro et al., 2010). We also saw a non-significant increase during the first week of exposure, but the later association was much stronger. Additionally, this earlier study examined all cardiovascular outcomes whereas the later associations seen in our study could pertain to a subset of cardiovascular outcomes. The increased risk three weeks later could also indicate readmission for complications related to an earlier visit for the same person. Some literature has estimated that in the U.S., SCD-related readmission to a hospital could be as high as 27% within 30 days (Kumar et al., 2020).

We acknowledge several limitations to this study. First, we lacked individual-level information about whether repeat events occurred for the same patient within the month. At the same time, however, any repeat visit would only underscore the need to prevent an event in the first place. Second, although recorded information indicating a diagnosis of SCD and SCT were likely accurate, PDD diagnoses codes for recorded thalassemia-related outcomes may be less exact (Snyder et al., 2017). Thus, we should interpret these findings with caution. Moreover, we utilized hospitalization data because they were available for more years than ERV data; however, the outcomes examined were also severe enough to require overnight admission and care. Thus, we could have missed less serious events that may still impact daily life. Finally, we saw large heterogeneity within CZ-level analyses. About 5 or 6 of the CZs contained most of the cases we examined, particularly for SCD and SCT. Because of the small number of cases in some analyses, small differences meant wide swings in effect estimates. We did observe strong associations in CZs with very hot daytime summer temperatures, including East Contra Costa County and the Central Valley (not shown). Differences in access to health, acclimatization, and weatherized housing, including air conditioning, may explain heterogeneity among zones and may reveal the need for smaller, more focused cohort studies per region.

Our study also contains several strengths. It is one of the first to explore hemoglobinopathy types and carrier status with hospitalization records spanning two-decades. Although we used state-wide records, our double-pronged approach did enable an initial regional analysis. The first stage results with large heterogeneity nonetheless informed us of a need for more focused study designs in the future. We are also one of the first to examine categories and both primary and secondary diagnoses of hemoglobinopathy and associations of heat. Because of this distinction between primary and secondary hospital diagnoses, we detected both acute and more lagged associations dependent on category and diagnosis level (primary or secondary) that may have eluded those just examining overall counts of hemoglobinopathies or doing an analysis over very short exposure periods. Discussed further below, we are also one of the first to consider in detail how climate, historical events, demographic changes, and structural barriers have and will continue to collectively influence U.S. hemoglobinopathy cases.

### 4.2 Hemoglobinopathy history and changing demographics in the U.S

Discussions on hemoglobinopathies in the U.S. should consider demographic factors, societal shifts, and a dearth in research that has severely neglected a large portion of the nation’s population. Climate change already affects minority populations disproportionately (Morello-Frosch and Obasogie, 2023), and the interaction between hemoglobinopathies and heat adds another layer of vulnerability for some. In addition to health concerns associated with a blood disorder or being a carrier, lack of funding for research, stigma associated with a genetic disease, and racism associated with treatment can compound complications associated with receiving adequate treatment (Farooq et al., 2020; Morello-Frosch and Obasogie, 2023; Naik and Haywood, 2015). Globally, SCD and thalassemia can affect a range of populations generally living in areas where malaria is endemic, particularly Africa, South Asia, the Middle East, and the Mediterranean (Bender et al., 2020) (Figure 7). Some reports estimate thalassemia carriers at 1.5% for βT and 5% for αT, respectively, of the global population (Kattamis et al., 2022).

**Figure 7.**
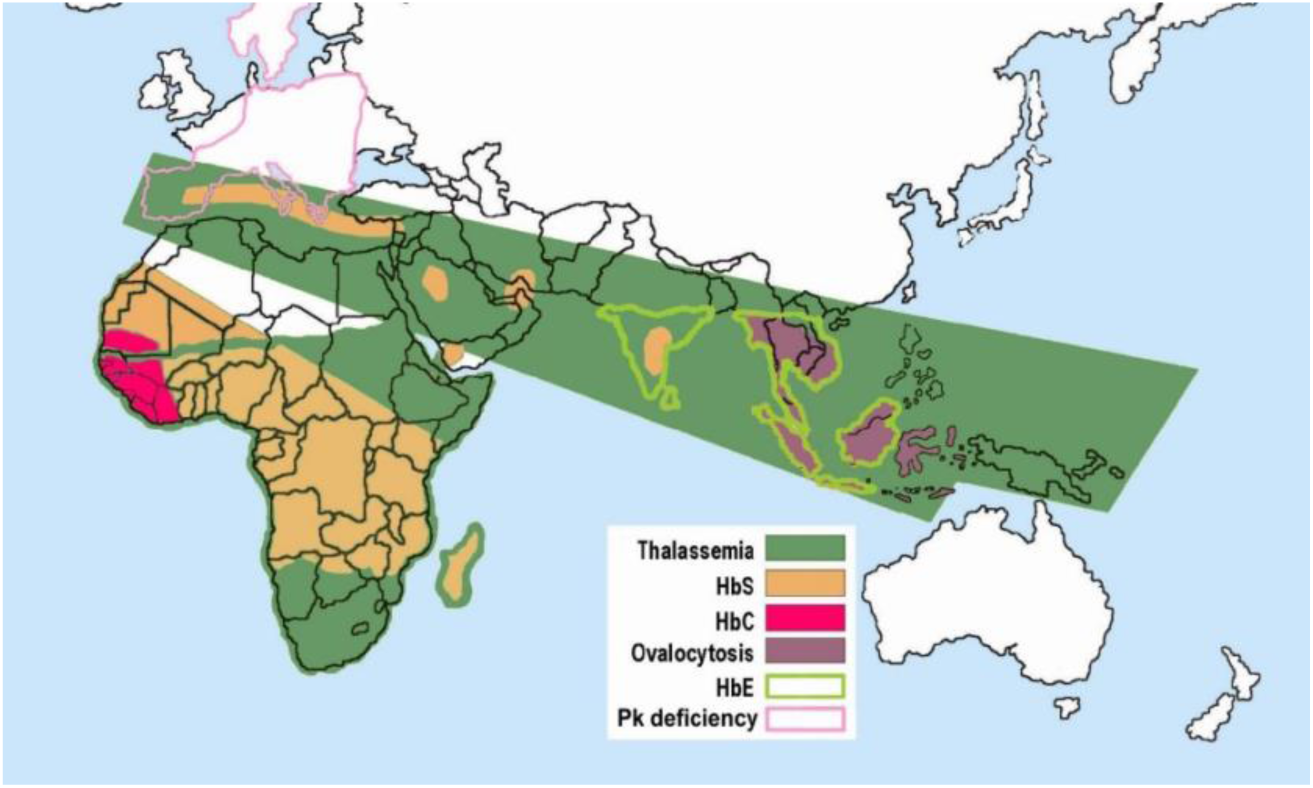
hemoglobinopathy Global Distribution compared to Malaria prevalence Source: Wikimedia Commons, Public Domain original artwork “Description Malaria versus sickle-cell trait distribution” by Armando Moreno Vranich

At the same time, however, SCD/SCT and thalassemia prevalence in the U.S. generally stems from the trans-Atlantic slave trade (Power-Hays and McGann, 2020) as well as some of the country’s historic racially and ethnically-motivated immigration policies that stymied immigration from Southern European, Middle Eastern, African, and Asian countries (United States Department of State). The Immigration Act of 1924 excluded immigration from all Asian countries and issued a quota systems that restricted immigration from Southern and Eastern Europe (United States Department of State). These new restrictions followed earlier legislation, such as the Chinese Exclusion Act of 1882 and various laws from 1790 to 1870 that excluded either the right to immigrate or citizenship to those of Asian heritage (United States Department of State).

Coincidentally, genetic predisposition to some autosomal recessive hemoglobinopathies is also more common among populations that originate from those restricted regions. Thus, although globally SCD and SCT can affect multiple populations, in the U.S., because of historic events and subsequent policies, those who identify as Black make up most of the patients with hemoglobinopathies, particularly SCD and SCT. Researchers in a recent study based in Michigan estimated that over 80 percent of those examined in the study with SCT or Sickle Cell Anemia were Black (Reeves et al., 2019). It should be noted, however, that anyone of any self-identified race/ethnicity could be a genetic carrier of a hemoglobinopathy, especially in a country as diverse as the U.S., because race is a complicated social construct and not based on a person’s full knowledge of their ancestry or national origins.

Although experts estimate at least 7% of Black Americans carry SCT, up to 3 million or approximately 1%, of the 2010 U.S. population are carriers from all self-identified race/ethnicity groups (Reeves et al., 2019). Thalassemia, on the other hand, involves more varied U.S. population groups, but the disease only gained national attention early in the Twentieth Century when newly-immigrant Italian children showed symptoms ((ASH), 2008). Along with a lack of sophisticated public health tracking among isolated immigrant and minority populations, who often faced segregation and marginalization ((CHSF), 2023), immigration restrictions on populations originating from high prevalence areas likely kept case numbers low for decades.

Recently, however, legislation has eased restrictions, and more equitable policies have increased immigration from numerous countries. Currently, most immigration stems from Asia and Latin America compared to 1960 when foreign-born populations from Asia, Mexico, and other Latin American countries made up just 4, 6, and 3 percent, respectively (Budiman, 2020). In contrast, the 2018 numbers for those residing in the U.S. who were born in the same three areas was 28, 25, and 25 percent (Budiman, 2020). In addition to new immigration, people identifying as multi-ethnic have increased, thus further blurring lines among populations with historically high prevalence (Jones N, 2021; Valle et al., 2022).

In addition to the disproportionate burden of disease at face level, some populations, particularly those identifying as Black, also confront challenges from structural barriers, such as inadequate treatment and access to medical care, medical debt, and food deserts that lead to poor nutrition and overall health. Historical redlining in areas riddled with concrete surfaces and lacking greenspace, poorly weatherized housing, and gentrification (which increases rent and housing insecurity) further exacerbate poor health by increasing temperature exposure level (Piel et al., 2017; Power-Hays and McGann, 2020; Reeves et al., 2022). In California, housing insecurity is especially high among those who identify as Black. A recent California governmental report found that despite comprising less than 6 percent of the state’s population, those identifying as Black make up over a quarter of the state’s unhoused population (Davalos, 2023). Moreover, Black Americans also share a disproportionate burden of the nation’s medical debt. A national report recently highlighted that among families having difficulty paying medical bills, those identifying as non-Hispanic Black represented the largest group among those who were surveyed (Cohen RA, 2023). These barriers, collectively, may worsen poor health outcomes, even among those who have a carrier condition that, on its own, is manageable.

Immigrant groups at higher risk of hemoglobinopathy-related complications face different barriers. Some individuals may never have received a proper diagnosis in their home countries (Kattamis et al., 2020). Even if aware, language barriers and lack of proper training among medical practitioners about prevalence in new immigrant groups may prevent easy access to information (Vichinsky et al., 2005). Considering occupational risk, some of these new residents may work in commercial kitchens, warehouses, and outdoors, conducting tasks that may put them at higher risk of dehydration or heat stroke. Many are farmworkers from Latin American countries with diverse backgrounds, and others (Hmong and ethnic Chinese) originate from areas where thalassemia is more prevalent (Apidechkul et al., 2021). Although knowledge about the diversity of high-risk populations is spreading, some healthcare providers may lack updated knowledge.

### 4.3 California Climate Commitment and Resolutions to Embed Equity in Research Efforts

Those with SCD and SCT as well as those working to provide care and conduct research in hemoglobinopathy communities likely already understand the connection between heat and hemoglobinopathies. The general population, however, may lack awareness that those with SCD and SCT have an increased risk of dehydration, heat stroke, and sickle cell-related complications because of extreme heat. In fact, even health care providers who do not practice in locales where SCD and SCT are more common may miss critical signs of a serious sickle cell-related complication in a population. Teachers, coaches, and others who work with juvenile populations who engage in strenuous activities may also lack information and training ((NATA), 2007). Furthermore, even if they are aware, these individuals or the public in general may not understand the complicated demographic changes altering historic prevalence patterns among populations.

Additionally, recent studies lay bare the reality of inequities those with SCD experience. Some estimate SCD patients live, on average, almost two decades fewer than those without (Jiao et al., 2023; Lubeck et al., 2019). Studies looking at SCT or other blood disorders and life expectancy are lacking. Others have noted less funding, both private and governmental, available to SCD and other rare blood diseases, despite SCD being identified as the most common genetic disorder in the U.S. In contrast, cystic fibrosis (CF), more common among those of Northern European heritage, receives more private and about the same federal funding, despite having a third of the prevalence (Farooq et al., 2020; Power-Hays and McGann, 2020). In fact, the last 10 federally funded, comprehensive care centers specifically for SCD closed in 2008 (Farooq et al., 2020; Grosse et al., 2009). Meanwhile, more than 100 centers, mostly funded privately, exist for CF. Historical racism associated with SCD, lack of private funding, and poorly centralized organization drive this inequity, but more government funding can close the gap (Farooq et al., 2020). Researchers also emphasize despite those under 18 having received improved care over the last couple of decades, adult SCD patients often vanish from primary care medical systems. Many such patients rely on emergency care as a source of primary care for their symptoms, resulting in subpar care management (Farooq et al., 2020). Given the potential for complications in later life, older SCD patients should receive primary care outside of ERVs. Similarly, officials should consider allocating additional funds for carrier studies, which could garner more robust evidence for heat and other exposure associations. With new technology bringing a cure for SCD to the forefront, ensuring people receive proper diagnosis and organized primary care throughout their lives will ensure fewer complications (Kolata, 2023).

To counteract the described structural inequities, newly developed state resolutions, such as A California for All, the California Strategic Growth Council’s Racial Equity Resolution and Equity Executive Order, EO N-16-22, make equity considerations in research a priority, particularly those related to climate change. With these thoughts in mind, the following points may inform policy and help alleviate disproportionate burdens not only in California but also other states, given nationwide historical structural inequalities and comorbidities unevenly affect some populations:

- Explain to the general population and policy makers the possible relationship between extreme heat and morbidity related to hemoglobinopathies, why climate change may increase morbidity, and who is at risk (young athletes and older individuals with multiple comorbidities who may be carriers).
- Recognize that although people of any race/ethnicity background may have a hemoglobinopathy, incidence of SCD or SCT is highest among those identifying as Black. Because of existing structural inequities, Black Americans face barriers to receiving adequate care.
- Inform the general population, health care providers, and policy makers that the demographics of blood disorders are changing, and that self-knowledge about SCD, SCT, and thalassemia may be lacking among newly-immigrant populations.
- Keeping in mind fair and efficient allocation of resources to build climate resiliency: provide additional resources for hospitals where heat-related visits could increase; provide funds for preventative measures, including providing for building resiliency and air conditioning in urban areas; require that employers provide shade, additional rest time, and healthcare checks for workers.
- Educate farm, factory, and warehouse workers who may engage in strenuous or outdoor activities about preventing morbidity related to their carrier status, such as easing into strenuous activity routines and drinking plenty of liquids. Inform policy makers and healthcare providers that heat exhaustion can occur in non-outdoor settings.
- Educate the public, health care providers, policy makers, and others about the stigma and controversy around sickle cell testing and take measures to ensure discrimination does not occur because of someone’s SCT status. Although knowing their health status is important for the individual to make informed decisions, others should not use this information to preclude anyone from participating in elite athletics or specific kinds of employment.
- Recognize other climate change-associated exposures (extreme temperature and diurnal temperature change, unseasonably cold weather, wind strength) may also present challenges, especially among unhoused populations, those in inadequate housing, or outdoor workers.
- Allocate funding to sickle cell care centers or find ways for SCD patients to find care outside of emergency rooms, especially after they turn 18. Help organizations become more centralized and provide support as they learn to navigate fundraising efforts. Provide funding for research, particularly for long-term studies involving carriers, as a gap measure.
- Provide translations of any guidelines or factsheets in numerous languages, especially as new treatments become available.

## 5. Conclusion

We observed an increased risk pattern of hemoglobinopathy-related health problems in California, where climate historically tends to be milder, following increased exposure to heat. Given the lack of research into heat and SCT in the general population, as opposed to military recruits or athletes, we believe longer term studies among those with SCT could reveal a worsening and increasing pattern of chronic complications related to heat not apparent in the short term. Similarly, disentangling differences between α and β thalassemia as well as the number of genetic carrier genes through focused studies could clarify exposure/outcome dynamics. Particularly, researchers should consider that older individuals with SCT or TM, who may have multiple comorbidities or organ damage, could face more immediate effects from heat whereas those who are younger may not. Because quality of care, occupation, acclimatization to heat, housing quality, and income could vary from place to place, regionally focused cohort studies would best elucidate true associations between heat exposure and Hemoglobinopathies and identify modifiable risk. With more knowledge, compassion, and understanding, policy makers can better allocate resources to a population who will otherwise face more negative impacts from climate change.

## Data Availability

The temperature data used in this study are available free to the public at: https://catalog.data.gov/dataset/aqs-data-mart; https://cimis.water.ca.gov/; http://www.ncdc.noaa.gov/oa/climate/climateresources.html
The health data used for this study if from the California Department of Health Care Access and Information (HCAI). Due to their sensitive, identifiable nature, these data are not available to the public, and anyone who would like access to them should contact HCAI.

https://catalog.data.gov/dataset/aqs-data-mart

https://cimis.water.ca.gov/

http://www.ncdc.noaa.gov/oa/climate/climateresources.html

## DECLARATIONS

### Ethics approval and consent to participate

We obtained Institutional Review Board (IRB) approval from California’s Committee for the Protection of Human Subjects (CPHS) prior to starting this study, which used exempt, registry-only health data and had no contact with any subjects.

### Consent for publication

Not applicable.

### Availability of data and materials

The temperature data used in this study are available free to the public at: https://catalog.data.gov/dataset/aqs-data-mart; https://cimis.water.ca.gov/; http://www.ncdc.noaa.gov/oa/climate/climateresources.html

The health data used for this study if from the California Department of Health Care Access and Information (HCAI). Due to their sensitive, identifiable nature, these data are not available to the public, and anyone who would like access to them should contact HCAI.

### Competing interests

The authors declare that they have no known competing financial interests or personal relationships that could have appeared to influence the work reported in this paper.

## Funding

Nothing to declare.

## Author contributions

DP: conceptualization, methodology, data curation and preparation, research and literature review, analysis, writing, figure and table preparation, editing, reviewing; BW: providing expertise on hemoglobinopathies, reviewing, and editing; KE: statistical and methodological advising and supervision, geospatial and map figure preparation, draft preparation, reviewing and editing; All authors commented on previous versions of the manuscript and read and approved the final manuscript.

## Acknowledgements

1. D. Pearson worked at OEHHA until April 2024 and completed the analysis for this project during that time. The authors would like to thank Brian Malig, MPH, for developing data curation methods for creating climatic and exposure datasets that we have adapted to complete this project, and Annie Chen, PhD, for her comments on project details and programming advice early on. We also gratefully acknowledge Craig Steinmaus, MD, Leona Scanlan, PhD, and Elaine Khan, PhD for reviewing this manuscript and providing valuable internal commentary on its content. Finally, we especially thank Susan Paulukonis, MPH, sharing her extensive knowledge of hemoglobinopathies, for going over early drafts of this manuscript, providing both expert and editorial comments, and providing other guidance as needed.

The opinions expressed in this article are those of the authors and do not represent those of the California Environmental Protection Agency, the Office of Environmental Health Hazard Assessment, the State of California, or the Governor’s Office.

## Notes

### Competing Interest Statement

The authors have declared no competing interest.

### Funding Statement

This study did not receive any funding

### Author Declarations

We obtained Institutional Review Board (IRB) approval from California Committee for the Protection of Human Subjects (CPHS) prior to starting this study that used exempt registry-only health data and had no contact with any subjects.

